# In Support of a Patient Navigator Program for Diabetic Foot Ulcer Care: A Qualitative Thematic Analysis and Pilot Studies

**DOI:** 10.1101/2025.11.25.25340998

**Authors:** Rebekah N. Williams, Jesica M. Flores, Kristin Harrington, Aliza Morani, Tejaswini Agarwal, Cassange Bitere, Walkiria Zamudio-Coronado, Ina Flores, Elizabeth C. Rhodes, Natalie Meriwether, JoAnna L. Hillman, Maya Fayfman, Marcos C. Schechter

**Author notes:** RNW and JMF contributed equally. JLH, MF, and MCS contributed equally.

## Abstract

**Objective:** The primary objective of this study was to inform and assist in planning the design of a patient navigator randomized controlled trial. We explored patient perceptions and preferences for the proposed intervention and their broader care experiences with diabetic foot ulcer (DFU) care. We additionally conducted a retrospective chart review to investigate if patient navigator phone calls are associated with increased clinic attendance and a pilot study of a 30-day post discharge navigator program for people with DFU.

**Methods:** This qualitative study involved two semi-structured focus group discussions (FDGs) with a total of 13 English-speaking patients with DFUs. Participants were recruited from an urban hospital in Atlanta (Grady Memorial Hospital). A deductive thematic analysis was performed on the discussion transcripts using a codebook structured around 20 prioritized constructs from the Consolidated Framework for Implementation Research (CFIR). We reviewed medical records for patients hospitalized with a DFU at Grady Memorial Hospital between January 2016 and December 2022 and compared post-hospital discharge DFU-related outpatient clinic attendance rates stratified by receipt or no receipt of a patient navigator phone call. The pilot study was a DFU navigator program with a primary outcome of 30-day post-hospital discharge clinic attendance (n=12).

**Results:** The thematic analysis revealed two primary domains shaping the patient experience: “Communication Gaps and Inconsistency” and “System Fatigue and Erosion of Trust.” The overall theme reflected a significant loss of confidence, with participants expressing skepticism and concerns that care sometimes seemed more focused on financial gain than on patient well-being. From these experiences, participants articulated a clear desire for a patient navigator who serves to advocate, coordinate, educate, and provide emotional support. Among 1285 patients hospitalized with a DFU, 153 (12%) received a post-hospital discharge patient navigator phone call. The 30-day post-hospital discharge DFU-related outpatient clinic attendance rate among those that did and did not receive a patient navigator phone call was 90% and 50%, respectively (p<.01). In the pilot navigator program, all 12 participants attended a DFU-related outpatient visit within 30 days of hospital discharge.

**Conclusion:** The proposed intervention of a patient navigation program directly addresses the foundational issues raised by participants of mistrust and fragmentation in DFU care. The findings indicate that trust-building, transparency, and reliable care coordination are essential requirements for successful patient navigation programs and, ultimately, for improved clinical outcomes in DFU care. The retrospective study and the pilot study demonstrate that high rates of outpatient clinic attendance are possible.

## 1. Introduction

More than 37 million Americans suffer from diabetes, where approximately 34% will develop a diabetic foot ulcer (DFU) in their lifetime and 1.6 million are affected by a DFU each year. DFUs lead to 80% of lower limb amputations and can increase the risk of death, as the mortality rate with ulcers is ∼30% and increases to over 70% following a major amputation.

Different populations within the United States are disproportionately impacted by DFU and subsequent amputations (Armstrong et al., 2023). Despite national hospitalization and survey data indicating improvements in diabetes care, minor and major amputation rates due to DFUs have increased (Geiss et al., 2019).

The management of diabetic foot ulcers has a significant effect on quality of life, as nutritional deficits, and depression are common in this population and associated with increased mortality (Armstrong et al., 2023). For instance, a qualitative study on the perceptions of diet quality in interviewed patients with DFUs reported a negative relationship with portion control, barriers to fresh, nutritious food due to limited mobility, and a lack of self-management knowledge stemming from mistrust in health professionals; many participants were unaware of the severity and possibility of amputation but preferred a personalized care intervention to support their dietary and DFU complications (Donnelly et al., 2022).

Care coordination among people with DFUs and/or amputations remains ineffectual despite the high risk of complications (Schechter et al., 2020). A retrospective study investigating outpatient clinic attendance and outcomes for DFU hospitalizations, albeit with limited data available, found only 53% attended post-discharge clinic appointments. Despite the known benefits of post-discharge DFU care, people with less severe presentations had lower attendance to clinics post discharge. Subsequently,18% of patients underwent a major amputation or died within 12 months of discharge (Mahgoub et al., 2022). Similar challenges in DFU care were studied within the healthcare system, where researchers found barriers on various levels through interviews. Patients individually stated financial strain because of health literacy and inadequate insurance. On the systemic level, patients reported strained relationships with providers led to challenges in securing follow-up appointments, inadequate preventative foot care and education prior to DFU development, and delays in care following DFU onset (Tan et al., 2022). The researchers suggested that these systemic barriers to diabetic foot care could be mitigated through specialized healthcare providers to reduce amputation risk. Consistent with this, patients who received preventative podiatric care following DFU development had lower rates of amputation, and long-term management supported by multidisciplinary diabetic services was associated with reduced preventable amputations (Armstrong et al. 2023). These findings demonstrate the need for coordinated, patient-centered care that addresses barriers to engagement and recovery; patient navigator programs represent one such approach.

Patient navigators support patients and/or their family members with access to healthcare resources such as appointments, medication, health education, and patient advocacy. A study that investigated patient navigators found four key domains of the role: navigating, facilitating, maintaining, and documenting. Navigators often conducted tasks such as identifying and mitigating challenges to attaining care, ensuring appointment attendance, reviewing patient conditions via medical record systems, and interacting with providers, nonclinical staff, and social workers or family (Parker et al., 2010). A similar study, focusing on cancer patient navigation, developed several components of the navigator role including community resource outreach, patient empowerment, communication, education through prevention and health promotion (Willis, 2013). Patient navigation programs utilize navigators such as nurses or non-clinical staff to overcome barriers to quality of care for patients and reduce inequities in chronic disease management (Foppa et al., 2023). A review of 60 recent patient navigation programs (PNPs) revealed that satisfaction increased, and outcomes improved among patients in PNPs with assigned patient navigators, including adherence to medication, treatment completion, reduced appointment wait-times and cancellations, and valuable emotional support (Kokorelias et al., 2021).

There has been research on the impact of patient navigators on general diabetes management, but their role in DFU care specifically are not well-studied (Loskutova et al., 2016). This was demonstrated in a systematic review of the impact of multidisciplinary clinical teams on DFU outcomes, which only found 33 observational studies as no randomized clinical trials (RCTs) have been conducted. Teams consisted of physician leaders with other members as needed, addressing glycemic control, wound management, vascular disease, and infection, and 94% of studies reported a decrease in major amputations from DFUs with team involvement (Musuuza et al., 2020). Such promising results convey the need for RCTs to test how patient navigator programs may provide similar and improved results for underserved populations suffering from DFUs and/or amputations, as current research on PNPs focuses on general diabetes symptoms. For instance, a recent PNP was designed to benefit glycemic control, self-care, and knowledge about diabetes. The researchers reported a significant decrease in HbA1c levels and an increase in adherence to self-care and glycemic control, concluding that patient navigation can identify and address barriers in improving diabetes care (Foppa et al., 2023). Similarly, a prospective cohort study evaluated a telephone-based PNP where navigators connected patients to community programs, provider referrals, and encouragement. Following the program, the proportion of patients reporting feeling overwhelmed decreased, and 90% of patients reported they would use the program in the future (Loskutova et al., 2016). These results warrant further research into the potential benefits of patient navigation programs on DFU and amputation outcomes. A navigator program combining components of medical and community connection, especially at a safety-net hospital, would provide a well-rounded resource for patients to regulate both diabetes and emotional support. One cohort study did evaluate a PNP at a safety-net hospital for diabetes outpatient care and found an increase in scheduled appointments and decrease in HbA1c levels, but the lack of DFU and amputation representation among patient navigation program studies remains an issue (Horný et al., 2017).

The complex and unique nature of DFU care, including the need for frequent wound care, guidance from multiple specialists, and high emotional toll of the disease, underlies the need for a comprehensive navigator program. The results of our previous qualitative analysis conducted in 2020 exemplify this notion, as barriers to care and engagement such as mistrust in healthcare systems and need for emotional support were prominent themes within these underserved focus groups (Fayfman et al., 2020). A comprehensive intervention that supports patients not only with post-discharge care coordination and appointment scheduling, but also with navigating the U.S. healthcare system, including transportation barriers, health literacy needs, and emotional assistance, has the potential to significantly improve DFU care.

## 2. Methods

### 2.1 Description of the Focus Group Discussions

The purpose of the focus groups was to validate the planned approach for the design of a patient navigator program (RCT) to achieve wound healing within 20 weeks. The study team held preliminary meetings with stakeholders to scope the needs of the focus group discussions (FDGs). We developed a protocol and semi-structured discussion guide for the focus groups. The FGD guide was based on the following research question: What are the perceptions of patients on the planned approach for a patient navigator program (RCT) with a goal of wound healing within 20 weeks?

Two FGDs were held over two days with patients from a large, urban hospital within Atlanta, Georgia. Each FGD lasted approximately 1.5 hours. The FGDs were led by a facilitator, and audio was recorded. All recordings were stored securely in compliance with IRB standards. The first FGD had nine participants, and the second had four. Handouts were provided to participants to review, which provided an overview of the planned intervention. The semi-structured FGD guide was co-developed by the study team and included 20 questions within 5 sections. The first section inquired about general interest in the patient navigator program led by a certified diabetes educator (e.g. “How easy or difficult would it be for you to participate in this program?”). The second section examined perceptions of specific program aspects based on the information given by the moderator (e.g. “What specific services/support would you hope to receive from the patient navigator program?”). This provided a segue into the third section, which discussed the patients’ preferred structure of the program (e.g. “How frequently would you like to interact with a patient navigator?”). Impressions of the control group as part of the study were conferred as the fourth section (e.g. “Would you still be willing to participate as a member of the control group if not selected for the patient navigator group”), and the final section concluded the focus group discussion with closing questions (e.g. “What additional thoughts or suggestions do you have for the patient navigator program?”). The FGD Guide is available in the appendix.

### 2.2 FDG Data Management and Analyses

All FGDs were transcribed by members of the study team using Microsoft Word. The dictate and transcribe functions of Microsoft Word were used for the transcription. Two research analysts each transcribed one of the FGDs, and any indecipherable dialogue was reexamined and verified by the facilitator.

The Consolidated Framework for Implementation Research <u>(CFIR)</u> theory was utilized to build a codebook that addressed patient perspectives on feasibility and motivations and the environmental and social constructs around implementation of an intervention. Following an initial review of the focus group transcripts’ data, 20 of 48 total CFIR constructs within five domains were selected that aligned with emerging themes. We grounded this approach based on recommendations from the CFIR User Guide and Coding Guidelines with the aim of emphasizing relevance and variation in the codebook. Two research analysts independently and blindly coded both transcripts using this codebook in MaxQDA. After coding the first transcript, the analysts and a supervisor met multiple times to adapt any needed changes to the codebook by consensus before proceeding to code the second transcript. Approximately 3-4 responses were labeled as crosstalk or inaudible due to inconsistent audio quality and were not included in coding. During thematic analysis, the team created a matrix of emergent themes defined by two or more codes along with exemplary quotes. To refine these thematic definitions and nuances, the two analysts and supervisor met frequently to review the codes and agree on consistency of themes across both coders.

### 2.3 Retrospective chart review and pilot studies

To complement the focus group discussions and obtain a signal on patient navigation effectiveness to improve outpatient clinic attendance in this population, we performed a retrospective chart of all patients hospitalized with a DFU at Grady Memorial Hospital between Jan 1, 2016, and Dec 31, 2022 and stratified DFU-related post-hospital discharge clinic attendance by receipt or no receipt of a patient navigator phone call. For univariate comparisons, differences in nominal variables were tested using either a Fisher’s exact or χ2 test and for continuous variables, either a Mann-Whitney or 2-sample t test was used as appropriate. A two-sided P value < .05 was considered significant.

We also conducted a concurrent 30-day pilot study of a navigator program to link patients with DFU care following discharge. In December 2023, we enrolled a separate group of 12 participants hospitalized with DFUs at Grady Memorial Hospital and assigned them certified diabetes care and education specialists (CCES) who conducted weekly calls for 4 weeks. These calls consisted of diabetes management, outpatient care coordination, appointment reminders, and addressing DFU-related concerns. All participants had a history of DFU or prior amputation.

The study was approved by the Emory Investigational Review Board (IRB) and the Grady Memorial Hospital Research Oversight Committee.

## 3. Results

### 3.1 FGD Participant Characteristics

English-speaking patients at the Grady Health System Diabetes Clinic who have a diagnosis of Diabetes Mellitus and have a diabetic foot ulcer with or without an amputation. Of the participants enrolled, mean age was 62 years-old (interquartile range 59-68 years old), 6 were female, and all identified non-Hispanic Black.

### 3.2 Domain 1. Communication

#### 3.2.1. Communication as the core of trust and engagement

Participants express a need for transparent, respectful and plain-language communication with the providers. They feel the providers do not explain the healing procedures such as wound debridement, leaving patients uninformed about the treatment they are getting. Patients express a need to be heard, have their questions answered, and be able to talk comfortably about various aspects of their condition.

> *“Has any doctor ever explained how healing happens? … No.” - Transcript 2*

> *“Just put it plain and let me know what I really need to do to make things better.” - Transcript 2*

> *“No, it’s no communication to you about how long this is going to be. Or how? What can you do besides just, you know, you know this will help you. You understand instead of the same old thing and sometimes you don’t want to come to the appointment.” - Transcript 1*

> *“Yeah, sometimes the doctors need to interact more with the patient, you know? I mean, not just, like, take a look at the foot, come back, scrape it, then you know, bandage it back up and like OK, see you in the next so or so time, you know. I mean, a little more information on what’s going on with your foot, you know, OK, what’s going on right there? Why is it still, you know, why is it still leaking? Why, you know, every time you come here and you scrape it, it bleeds, you know? I mean, it never bleeds, not before I came here, but as soon as you start scraping it starts bleeding, so going back to square one. - Transcript 1*

#### 3.2.2. Communication is an emotional experience

Patients consistently emphasize the need to be heard, listened to, and cared for. When they are not given proper information about the technical processes, a patient is left not understanding if they are getting better or worse. They feel they are going back to square one, furthering increasing the mistrust. This also leaves them unable to advocate for their own bodily/ medical condition. Patients say that lapses in communication are routine and describe it as an expected part of care. They also feel dismissed when providers are not concerned about their needs, reinforcing the power dynamics at play between the provider and patient.

> *“I’m going to be honest with you. Communication is the key. Showing them really, that you really care versus just saying something. If I’m in a hospital and I don’t know all your codes and stuff like that and I’m about to die, tell me I’m about to die. I want to be in the name this turn.” - Transcript 1*

> *“So it’s just more communication. Just comment. Just, you know, like I said, just look, look at it, scrape it, clean it up as I thought they said they clean it up. But that cleaning up, puts you back to square 1.” - Transcript 2*

> *“You as my provider should be like, OK, this is what you need to do… let me know those things.” - Transcript 2*

### 3.3. Domain 2: System fatigue and erosion of trust

#### 3.3.1. Mistrust in the healthcare system/ providers

Patients are skeptical about the care they are receiving; they express their frustration with repeated procedures and express concerns about whether it is medically necessary or about money.

> *“Stop using me as a guinea pig… you’re going to keep scraping it so I can keep coming back so you can grab a salary.” - Transcript 2*

> *“They get more money, more treatment when they cut all the toes off… I didn’t have to get all my toes cut off. But it’s done now.” - Transcript 2*

> *“But oh. Alright, let me not make you mad. You understand I’m not calling no name, but that all of you just scrape, scrape, scrape, scrape, scrape and then you start to bleed and then we wrap you up. You understand?” - Transcript 1*

> *“. Well, I’m not saying this gonna heal it. Because sometimes they say they’re going, they’re going to take care of the problem. And then when they don’t, you live right there. So the going through this 20 weeks, I don’t want to be. Like I said, I don’t want to be used as a Guinea pig. I don’t want to be used to try to say, well, we’re going to try this method on you. And with that method on you. And then it still don’t work. You know, for the next person. It ain’t all about just me. It ain’t all about just him, him or her. It’s for the next person.” - Transcript 1*

Patients express that sometimes a provider is not well-versed with the patient’s history, and their treatment differs from another provider who the patient was previously seeing. There may also be poor communication among different providers, which can lead to confusion, and ultimately, mistrust.

> *“I had one doctor telling me one thing, then another doctor coming in and telling me something else.” - Transcript 2*

> *“Too many doctors coming here saying one thing… they’re not communicating.” - Transcript 2*

### 3.4. Domain 3: Desire for patient centered and compassionate care

#### 3.4.1. Patients express a need for patient-centered care

Patients express a need to be heard, respected, and emotionally supported. They value a provider who is calm, genuine, non-judgmental, and not only treats the wound but also treats the person. They need a provider who will talk to them patiently about their issues and understand them.

> *“Not hot tempered… calm, genuine, not making me feel incompetent.” - Transcript 2*

> *“Someone who you can sit and talk to… a doctor visit is in and out. I’ll say something, but I don’t get to really, you know, elaborate more with the doctor, you know, someone who will understand and can give you more detail I guess.” - Transcript 2*

> *“He don’t make you feel like what you’re saying is not a factor… he’s very tentative.”- Transcript 2*

#### 3.4.2. Compassion is seen through gestures and not just words

Patients express that compassion is felt through time, tone and small gestures of humanity like being attentive, asking about family and not rushing with the appointment. It is about micro-interactions and not grand acts. Patients interpret these behaviors as evidence that they matter.

> *“If a person comes in [doctor], you’re sitting down, everybody’s talking. The other person starts looking at their watch, yeah, that pisses a person off, you understand? … seems like you got something else to do that’s more important than what I’m trying to say, you know.” - Transcript 2*

> *“You know, he’s very good, very good on, he even shows concern about things that it’s not even related to. Just basically actually just how your day went and how, how you feeling? How’s the family? Stuff like that. That’s what makes you feel good; it makes you want to open up to that person.” - Transcript 1*

### 3.5. Domain 4: Healing through connection: Peers, faith and shared experiences

#### 3.5.1. Value of relationships and community support

Participants highlight the strength found in shared experiences, small groups, and peer contexts. They express that these platforms provide them with a chance to talk to people going through the same situation, and they learn from each other’s experiences. Participants seek comfort, solidarity, and confidentiality in each other’s company. Participants desire the opportunity to have informal meet ups such as group exercise programs. Small groups suggest better motivation to join and stay.

> *“I’m gaining strength from them, even though I don’t know none of y’all.” - Transcript 2*

> *“Being comfortable being able to share… you made it comfortable for us.”- Transcript 2*

> *“I mean also the the exercise as far as the gym, you know? All that costs funds. Umm Maybe they could get a contract with such and such and such, or for the people. To be able to say, OK, well, let’s let’s get together and let’s go to the gym today. Let’s go to. Then let’s go to. The OK, let’s go to the big pool” - Transcript 1*

> *where we can all go in there and swim, you know. And I think all that stuff there would just enhance you, man. It would make you feel better.” - Transcript 1*

#### 3.5.2. Faith and Spiritual coping

Participants express deep faith in God. They highlight that they gain strength knowing whatever is happening is because of God and he is there to take care of them. Spirituality seems to play an important role in resilience where other methods fail. Faith sustains when medical treatment fails or leaves a gap.

> *“If God gave me certain testimonies and certain things to share with people, why sit down over it? Why not share it? And then it’s not just about no faith-based church. I have a good faith, faith church. I’m able, but it’s about each one teach one.”- Transcript 2*

> *“I ain’t worried about all that to the bottom? That going to come? Because if you believe in God, it’s already taken care of.”- Transcript 1*

> *“So if God. He gives doctors certain knowledge to knowing how to cut and do all that. I believe sometimes the doctor sees what they see. But the thing is though, my whole healing is through the grace of God, that’s through my healing now.”- Transcript 2*

### 3.6. Domain 5: Holistic Autonomy through knowledge

#### 3.6.1. Need for education

Patients want to be informed and educated about their condition and the treatment they receive. They want to understand why certain procedures are being done, what medications they are taking, and the impact all this is having on their health.

> *“Yeah, sometimes the doctors need to interact more with the patient, you know? I mean, not just, like, take a look at the foot, come back, scrape it, then you know, bandage it back up and like OK, see you in the next so or so time, you know. I mean, a little more information on what’s going on with your foot, you know, OK, what’s going on right there? Why is it still, you know, why is it still leaking? Why, you know, every time you come here and you scrape it, it bleeds, you know? I mean, it never bleeds, not before I came here, but as soon as you start scraping it starts bleeding, so going back to square one.”-Transcript 1*

> *“Tell me what I’m putting in my body… I want to know what I’m taking.” - Transcript 2*

#### 3.6.2. Knowledge as a tool for self-efficacy

The participants emphasize that education is an important component in them being able to take control of their condition. They want to have rightful ownership for health information. Knowledge restores agency in a system that often strips it away. Being informed is seen as essential for self-efficacy and decision making.

> *“I think that’s one of the benefits for me would come out of, you know, just being made aware of, hey, we can control this. We can get this under control and that’s all a part of the education part of you know and it keeps you focused on (…) something other than you know” - Transcript 1*

> *“P1: Well, I think and I’ll go back to it will almost everybody has said in his education, the more you are educated on what is wrong with you and how you can better treat it, the better off we are going to be, you know, and not just.And I think the other thing is just getting getting being able to get the treatment that we need.” - Transcript 1*

### 3.7. Domain 6: Patients anticipated outcomes from the program

#### 3.7.1. Expectations from the navigator

The patient navigator is seen as a bridge connecting the patients with the healthcare system. They are viewed as someone to help patients feel “seen” and “heard” between visits. Some desired traits of the navigator include knowledgeable (about diabetes, procedures, and medications), attentive, communicative, listener, consistent, compassionate, non-judgmental, calm, and available. An ideal navigator should be able to guide the patients through the procedures and be available when the doctor cannot be reached.

Additionally, it would be helpful if the navigator would also help patients overcome logistical barriers like assisting them with transportation.

> *“Someone who you can sit and talk to, you know, cause sometimes a doctor visit is, you know, I mean, you get just like in and out I’ll say something, but I don’t get to really, you know, elaborate more with the doctor, you know, someone who will understand and can give you more detail I guess.”- Transcript 1*

> *“Someone who knows what they’re talking about… who can make it make sense.”- Transcript 2 “Calm, no attitude, not hot tempered.” - Transcript 2*

> *“The Navigator will also assist you with the riding back and forth to your appointment… I think that will be … very much … helpful.” - Transcript 1*

#### 3.7.2. Motivation to stay in the program

The patients emphasize clinical improvement as an important component to continue the program. As alluded to earlier, patients want to be educated about their condition and how they can take control of their care. Some patients talk about their experiences of similar programs where a lot of things were promised but not delivered, hence, they want to see consistency in the program delivery. Additionally, for some of them monetary incentives will be a motivation to join and stay.

> *“So I mean (.) I feel like the focus you put on is wound management, you know I’m saying because you know that’s the main problem.”- Transcript 1*

> *“The patients are motivated to join, however, they are not sure if the program will actually be consistent and provide them what they need. They want to be helpful that they will get real help” - Transcript 1*

> *“Umm In my case, as I indicated, and I’m not trying to sound like I’m hungry for the money, but yeah that that’s the motivation factor”- Transcript 1*

#### 3.7.3. Preferences for the program- modes of delivery, services offered, frequency

In terms of needs, people were flexible about certain services like mode of communication, where the patients are okay with either the navigator reaching out to them or them reaching out to the navigator. Some patients indicated that meeting the navigator once a week would be preferable while others emphasized that they want the navigator to be present when they need them. Some patients suggested a preference for meetings in conjunction with their regular clinic visit. In terms of the mode, most patients prefer in-person meetings while a few are happy with a phone call. Preferred visit duration ranged from 15 min to 1 hour.

> *“In terms of communication, both services would work- the navigator reaching out to them and them reaching out to the navigator “- Transcript 1*

> *“Instead of having it for that long availability, why not each time you visit you come to the clinic or at the doctor you someone is here to talk to.”-Transcript 1*

> *“The meeting can be either at their house or in the office”- Transcript 1*

### 3.8. Results of Retrospective Chart Review

Among 1285 patients hospitalized with a DFU at Grady Memorial Hospital between January 1, 2016 and December 31, 2022, 153 (12%) received a patient navigator phone call after hospital discharge. Patient characteristics are shown in Table 1. Patients that received a navigator call were younger, more likely to have a hemoglobin A1c ≥8.0%, and more likely to be uninsured compared to patients who did not receive a phone call. Patients who received a patient navigator call where significantly more likely to attend DFU-related visit compared to those who did not receive call at 7-day (39% vs 15%), 14-day (68% vs 32%), and 30-day (90% vs 50%) post-hospital discharge (p<.01 for all comparisons).

**Table 1:**
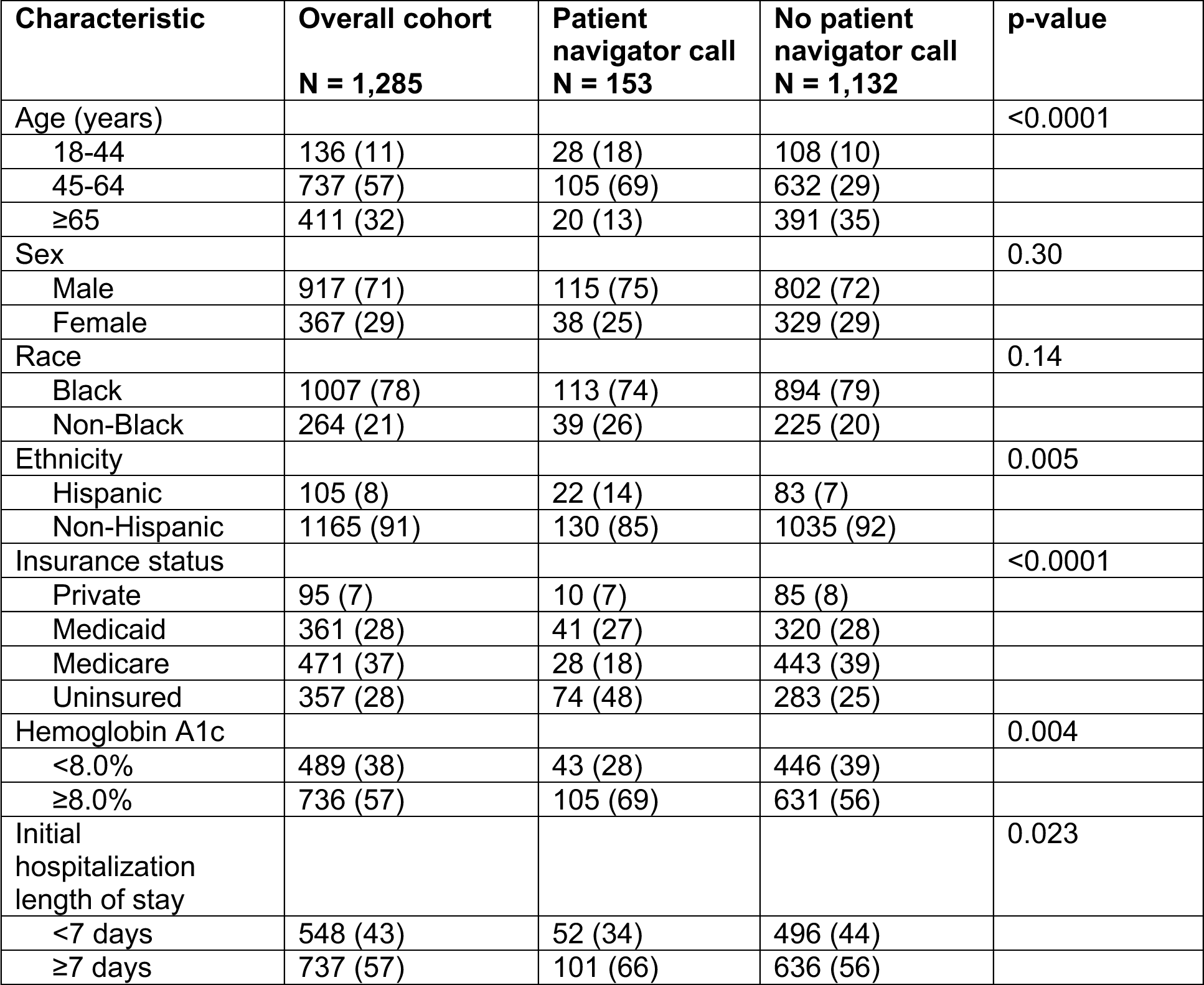
Comparison of baseline characteristics between patients that did and did not receive a patient navigator call within 30 days of hospital discharge.

### 3.9. Results of Pilot Navigator Program for post-hospitalization DFU care

We enrolled twelve participants in the program in December 2023. The median (IQR) age was 53 (42-59) years; 8 (73)% were male and 73% identified as non-Hispanic Black. The median (IQR) baseline HbA1c was 8.2% (7.7-11.1). Diabetes distress scores were high median (IQR) 2.9 (1.7-3.2). All participants attended a DFU-related outpatient visit within 30 days of hospital discharge. Ten of the 12 participants used a CDCES intervention aside from appointment reminders, including setting up transportation (n=1), wound care plan clarification (n=1), antibiotic adverse effect management (n=1), rescheduling appointments (n=3), severe hyper/hypoglycemia management (n=2), and insulin education and dose – modification (n=6).

## 4. Discussion

The findings from this study indicate a need to reconsider how barriers to (DFU) care are conceptualized, shifting attention from logistical challenges to a deeper erosion of trust. This section interprets the significance of these results, situates them within the existing body of research, and outlines their implications for clinical practice, health system design, and the preparation and integration of patient navigators into the healthcare system.

While previous research demonstrates that Patient Navigation Programs (PNPs) improve system-level outcomes such as appointment adherence, the findings of this study suggest that their value also lies in fostering interpersonal connection, providing advocacy, and helping to build trust between the patients, providers, and healthcare systems as a whole. Effective navigation extends beyond logistical coordination to include restoring confidence in the care relationship with a patient.

An important and distinctive theme was participants’ expression of financial doubt. The perception that common procedures may serve financial interests reflects a context-specific mistrust of the healthcare system. This financial concern represents a significant barrier to engagement, especially for underserved safety-net populations.

Health systems must invest in contextualized training for both providers and navigators in transparent, respectful, and plain-language communication, recognizing this as a core component of providing essential emotional support. To combat patient-provider mistrust, health care institutions should implement protocols that engage patients in understanding their disease, treatment rationale, anticipated recovery timelines, and expected costs.

To meet the complex needs identified by patients, navigators must be trained and integrated strategically. Training curricula must prioritize relational competencies over logistical ones, recognizing that the navigator’s primary function, as defined by patients, is to strengthen relationships and rebuild trust disrupted by systemic inconsistency. Structurally, navigators must be fully integrated into multidisciplinary care teams and empowered with the authority to review patient records and advocate for clarity and unified recommendations among members of the care team.

The study’s findings both align with and build upon previous research, introducing a deeper layer of relational necessity to the understanding of barriers in DFU care. The study reframes barriers to Diabetic Foot Ulcer (DFU) care, shifting the focus from logistics and coordination to a relational frame and breakdown in confidence caused by shortcomings common to many healthcare systems (i.e. limited provider time, fragmentation of care)(Tan et al., 2022). Key novel findings include skepticism among participants, who often perceive common procedures as unnecessary or excessive, which acts as a powerful limitation to engagement, leading to calls for increased financial and clinical transparency in care. Challenges such as care fragmentation cannot be addressed without significant financial investment, such as hospital wide multidisciplinary programs. A patient navigator may serve as a more cost-effective and easily implemented intervention to bridge the patient and providers to improve communication and patient self-efficacy. This study advocates for instituting and funding integrated PNPs that are knowledgeable about both disease and treatment and can serve as a mediary between the health care providers and patients.

The retrospective study suggested patient navigators can increase outpatient clinic attendance and this was corroborated in our pilot program. In our pilot, we connected dedicated CDCEs to each patient. Though multiple providers were involved in patient’s DFU care, continuity among the CDCEs may have served as a home base for participants to allow for effective engagement. The CDCES-led patient navigator intervention was associated with substantial improvement in 30 days post-discharge clinic attendance rates for patients hospitalized with DFUs and CDCES provided comprehensive care, addressing multiple aspects of diabetes and wound management.

### 4.1 Limitations

Throughout the process our researchers sought to maintain rigorous research methods, but there were areas of improvement in our design.

The small sample of 13 participants was recruited from a single safety-net hospital system. This context-specific sample limits the generalizability of the findings to other geographic or clinical settings

The audio quality of the FGDs was inconsistent in some areas of the transcripts, with significant background noise, multiple patients talking at once, or low microphone volume. The presence of these disruptors made it difficult to distinguish sentences, and approximately 3-4 responses were labeled as crosstalk or inaudible. This limitation may have influenced the scope of thematic analysis as some dialogue may have been lost, but transcripts were reviewed multiple times by the facilitator and research team for accuracy, and future research can consider multiple microphones to mitigate the issue.

Due to the deductive approach for the codebook, the researchers had differing approaches to applying the code definitions in the transcripts. To counter this, the team had frequent meetings to align on the overall emerging themes and how they were seen in the transcripts. Moving forward, we would recommend additional time to allow the researchers to learn and practice using the codebook if applying a deductive approach.

These limitations, while important, do not diminish the clarity of the core message emerging from the participants shared experiences, which points toward the study’s central conclusion.

## 5. Conclusion

In summary, patients in this study identified personalized support, trust, contextual understanding, and consistent communication as essential priorities for the design of effective patient navigation programs that are patient-centered, and sustainable. Streamlined communication between providers was also referred to as a concern for improving diabetes care. Building on this work, future research should focus on testing structured navigation interventions that explicitly incorporate these patient-informed elements. The impact of such interventions should be rigorously evaluated against both clinical outcomes, such as glycemic control, and key self-management outcomes, thereby connecting patient perspectives to tangible health improvements. Ultimately, implementing navigation programs that genuinely reflect patients stated needs and priorities may significantly enhance the quality and outcomes of diabetes care delivery.

## Supporting information

The FGD Guide is available in the appendix.

## Data Availability

All data produced in the present study are available upon reasonable request to the authors

## Grants

### Author Contributions

MF and MCS designed the study, reviewed the study design, and co-authored the manuscript. JF recruited participants. KWZC and IF conducted the pilot study. KH performed the retrospective study data analysis. JLH, MCS, and MF designed the FGD guide. JLH conducted the FGD sessions. AM and TA transcribed the FGD sessions, performed data entry, and analyzed the data with RNW. JLH, CB, RNW, TA, and AM wrote the draft of the manuscript. ECR, NW, and all authors edited the manuscript and approved its final version.

### Funding

This study was partially supported by the National Institute of Diabetes and Digestive and Kidney Diseases awards R01DK139326 (Fayfman and Schechter), and P30DK111024 (Fayfman and Schechter), in addition to an Emory Medical Care Foundation Programmatic Support (Schechter and Fayfman) and an Emory Department of Medicine FAME award (Schechter). MF is partially supported by National Institute of Diabetes and Digestive and Kidney Diseases K23DK124647 award.

