## Supplementary material for "In Support of a Patient Navigator Program for Diabetic Foot Ulcer Care: A Qualitative Thematic Analysis and Pilot Studies": The FGD Guide is available in the appendix.

**Grady Health System Focus Group Protocol**

**Patient Navigator (RCT), January 2024**

**RESEARCH QUESTION:** What are the perceptions of patients on the planned approach for a patient navigator program (RCT) with a goal of wound healing within 20 weeks?

**STUDY POPULATION:** English-speaking patients at the Grady Health System Diabetes Clinic who have a diagnosis of Diabetes Mellitus and have a diabetic foot ulcer with or without an amputation.

**Stage One: Greeting and Introduction (2 minutes)**

- Purpose: Welcome participants and express appreciation
- Things to include in welcome:
  - Introduction of yourself and your role
  - The purpose of today’s focus group
  - Signing of consent form

Sample Script: *Good afternoon, everyone. I would like to thank you all for coming today. I am JoAnna Hillman and I am partnering with Grady physicians to conduct and moderate this focus group*

**Stage Two: Utilities (3 minutes)**

- Purpose: Setting the stage for the session
- Things to include:
  - Confidentiality:
    - Highlight definition of confidentiality
  - Encourage turning off mobile phones and staying in the room.
  - Recording:
    - Highlight the presence of audio equipment
  - Note-taking
    - Explain that the moderator will take occasional notes to support the audio recording
    - Assure confidentiality

Sample Script: *These sessions are being recorded in order to gain the fullest information from the comments you make. The tapes will be transcribed and listen and read only in strict confidentiality. Your comments will be transcribed using a pseudonym (ie: participant 1, participant 2, etc). This information will only be used by those involved in this project.*

**Stage Three: Expectations (2 minutes)**

- Purpose: Establishing a safe environment
- Things to include:
  - Comfort: participants should feel free to speak openly and freely; You don't need to agree with others, but you must listen respectfully as others share their views
  - Use of “I” and “We”:
    - Participants can speak on their own experiences and preferences
    - Participants can also speak on the needs and preferences they have noticed of others around them
  - There are no right or wrong answers
  - Use of forward-thinking solutions are encouraged. While you may have had challenging experiences in the past navigating care, we are considering a program to improve this, so your input and recommendations today are critical to our program development.

**Stage Four: Icebreaker (8 minutes)**

Sample Script: *Let’s go around the circle and tell everyone your name, and <<ice-breaker question>>.*

- Allow chatter. Then quickly refocus without talking over anyone.

Sample Script: *The discussion will last about an hour. Please help yourselves to the refreshments provided. Are there any questions before we start? <<Indicate beginning the recording.>>*

**Stage Four: Program Description (5 minutes)**

Purpose: To provide a clear description of the planned patient navigator program to anchor the focus group discussion

Diabetic foot wounds are a common problem at Grady and often lead to amputations. We previously interviewed patients with diabetic foot wounds and learned about the challenges to foot wound care at Grady. We want to make it easier for patients to have good foot wound care at Grady and designed a program that we hope help patients heal their foot wounds.

This program will be led by a patient navigator. A patient navigator is a healthcare worker that is available to help guide you through the health care system and overcome the barriers that prevent you from the getting the care you need. In this program, the patient navigator will be a certified diabetes educator, meaning they can educate patients on how manage their diabetes and can help adjust diabetes medications.

We would like to test a 20-week program where you would have a call with a patient navigator once per week.

**Diabetes management**: the patient navigator will review your blood sugars and help adjust your diabetes medications. If your sugars are dangerously high or low, the patient navigator will reach out to a doctor.

**Wound management:** the patient navigator will review how you are doing wound care and make sure it is in line with what the doctors recommended. If desired, you can send pictures of your foot to the patient navigator. If there are concerns the wound is getting worse, the patient navigator will reach out to a doctor. The patient navigator will also help you get wound care supplies if needed.

**Antibiotic management:** if you are prescribed an antibiotic for your foot, the patient navigator will make sure you are taking it correctly. The patient navigator will reach out to a doctor if you develop antibiotic side effects.

**Care coordination:** diabetic foot wounds require many outpatient visits with different doctors. The patient navigator will make sure you know of all your appointments and try to help you reschedule appointments to better fit your availability.

**Transportation:** coming to so many outpatient visits requires transportation. The program will offer you 50 dollars’ worth of transportation support to use for appointments every 2 weeks for 20 weeks (500 dollars total). This can be in the form of rideshare, Grady parking vouchers, or gas gift cards. This is for transportation to and from Grady for your medical care only.

**Screening and resources:** many people struggle with issues such as depression, food insecurity, financial resource and housing instability, tobacco and alcohol use. This program will screen for these issues and provide resources and referrals, if needed.

**Stage Five: Discussion Questions (70 minutes)
KEY QUESTIONS**

A. **General interest in the patient navigator program led by a certified diabetes educator**

I would like to talk about your general interest in participating in a program like I’ve just described. Remember there are no right or wrong answers so feel free to tell us what you think.

1. Has anyone heard of patient navigators before? Has anyone ever worked with one? *Probe: impressions, experience*
2. What are your thoughts on participating in this patient navigator program?
   a. *If negative, probe for reasons for limited interest*
3. What are some factors that would impact your decision to participate? *Probe: trust of the medical system in comparison to a patient navigator, mitigation strategies for factors*
4. How easy or difficult would it be for you to participate in this program? *Probe: specific factors that might discourage individuals from seeking assistance from a patient navigato*r
5. What sorts of health benefits would you hope to see if you were to participate in this program?
B. **Perceptions of specific program aspects**

Now, I would like to know more about your impressions of the specific components of the program (review key components from description above).

1. Of all the things we've talked about, what component would have been most important to you, given your experience?
2. One of the program components will be supporting transportation needs to engage in the program. This could include parking validation, ride share arrangements, or a gas gift card. Would one of these options meet your needs? Would the amount be sufficient for your transportation needs?
3. What specific services or support would you hope to receive from the patient navigator program that we have not discussed? *Probe: emotional support, advocacy, attrition prevention*

Emotional support:

- Are there specific situations in your healthcare experience where emotional support would be particularly valuable?
- Probe: How important is it for a patient navigator program to involve and support your family or caregivers?
- What are your thoughts on the patient navigator program partnering with local community organizations and resources? *Probe: Suggestions for specific types of organizations, for which components*

C. **Preferred structure of Patient Navigator program**

Now, I would like to talk about how you would most like to see this program work.

1. Describe your ideal patient navigator. What characteristics would a patient navigator have that would be important to you?
2. How frequently would you like to interact with a patient navigator? *Probe: weekly, every other week*. How much time are you willing to commit to engaging with a patient navigator at this frequency?
3. How would you prefer to communicate with a patient navigator *(e.g., in-person, phone, video, email, text)? If in-person, probe for best location?*
4. How would you prefer to connect with a patient navigator; would you like them to reach out to you or you reach out to them?
5. What are your thoughts on including group sessions or peer support groups as part of a patient navigator program? *Probe: Meeting with those with lived experience, what would be wanted/appropriate here (Education, support, counseling)?*
6. What would motivate you to stay engaged throughout the patient navigator program? *Probe: How might other factors such as work or family commitments, impact your ability to stick with the program? Anything else that might cause you to drop out of the program?*
D. **Impressions of the control group**

As part of this project, there will be a group that will be part of the study and receive reminder phone calls, but not in the patient navigator program.

1. Would you still be willing to participate as a member of the control group if not selected for the patient navigator group?

**CLOSING QUESTIONS**

Finally, I’d like to be sure we’ve covered all your thoughts from today.

1. Based on any challenges you have faced navigating the healthcare system in the past, how do you believe a patient navigator program could benefit you or others in the community?
2. What additional thoughts or suggestions do you have for the patient navigator program?

**CONCLUSION**

We are now reaching the end of the discussion.

**Stage Six: Recap (3 min)**

- Review the main points of the discussion

**Stage Seven: Gratitude and Future of Data (2 minutes)**

- Purpose: Express appreciation
- Things to include:
  - Emphasize the importance of their voice
  - Dismiss participants with a big thank you!

Sample script: *“I would like to thank you all very much for your time and active participation in this discussion. Your perspective is greatly appreciated and has helped us understand your impressions of this planned program.”*

**Stage Eight: Wrap up**

- Purpose: Collect materials
  - Facilitator should ensure that all materials and recordings are collected and saved
